# Efficacy and Safety of Tregopil, a Novel, Ultra-Rapid Acting Oral Prandial Insulin Analog, as Part of a Basal-Bolus Regimen in Type 2 Diabetes: A Randomized, Active Controlled Phase 3 Study

**DOI:** 10.1101/2022.02.15.22270708

**Authors:** Harold E Lebovitz, Alexander Fleming, Alan D Cherrington, Shashank Joshi, Sandeep N. Athalye, Subramanian Loganathan, Ashwini Vishweswaramurthy, Jayanti Panda, Ashwani Marwah

## Abstract

**OBJECTIVE:** Efficacy and safety of ultra-rapid acting oral prandial insulin Tregopil (Tregopil) was compared with insulin aspart (IAsp) in patients with type-2 diabetes on stable doses of insulin glargine and metformin.

**RESEARCH DESIGN AND METHODS:** In this open-label, active-controlled trial, patients with type-2 diabetes, with HbA_1c_ ≥7% and ≤9% and 2-h postprandial glucose (PPG) ≥180 mg/dL were randomized (1:1:1) to Tregopil (30 mg [*n*=30], 45 mg [*n*=31]) and IAsp (*n*=30; dose titrated based on self-monitored blood glucose [SMBG]). Postprandial plasma glucose excursion (PPGE) and PPG were assessed from the standardized test meal (STM) and 9- point SMBG. The primary outcome measure was change from baseline (CFB) in HbA_1c_ at week 24.

**RESULTS:** The Tregopil (30 mg) arm showed significantly lesser 1-h PPGE (CFB) excursion after the STM versus IAsp (Estimated Treatment Difference [ETD], 95% CI, -45.33 mg/dL [-71.91, -8.75], *P*=0.001) and 1-h PPG trended towards a better control. The combined Tregopil group (30+45 mg) showed lower PPGE at 15 mins as compared to IAsp. Meal-wise analysis showed lower 1-h PPGE and PPG in the Tregopil groups post-breakfast. Clinically significant hypoglycemia was lower with Tregopil groups versus IAsp (rate ratio: 0.69).

**CONCLUSIONS:** Tregopil demonstrated an ultra-fast onset and short-duration prandial insulin profile with good safety. Tregopil improved the 1-h PPG and overall PPG control compared to IAsp. A further reduction in HbA_1c_ compared to baseline was not observed, likely as a result of variability in the control of fasting glucose level over the duration of the study.

## INTRODUCTION

Patients with type 2 diabetes who fail to achieve glycemic targets despite oral antidiabetic drugs (OADs) and lifestyle modifications require insulin therapy (1) and subcutaneous (SC) injection is the most commonly used route of administration (2). However, the SC injection is associated with challenges like needle phobia, injection site pain, lipodystrophy and may lead to patient noncompliance. Further, the occurrence of peripheral hyperinsulinemia (3), may result in adverse effects such as weight gain and hypoglycemia (4). Oral delivery of insulin has been long sought after. Insulin delivered orally is absorbed directly into the portal circulation, and resembles the pancreatic secretion of insulin and its portal transport, resulting in a higher hepatic insulin exposure (5). Hepatic delivery of oral insulin is associated with reduced risk of hypoglycemia (6), stabilized body weight (5), improved therapeutic compliance, and hence a better quality of life (7).

In patients with type 2 diabetes, postprandial glucose (PPG) levels typically peak about 2-h after a meal (8). Elevation in PPG is due to loss of first-phase insulin secretion, reduced insulin sensitivity in peripheral tissues, and decreased suppression of hepatic glucose output after meals (9). Bolus premeal insulin treatment reduces PPG excursion (PPGE) in patients with type 2 diabetes (10). Compared to the regular human insulin, first-generation, rapid-acting insulin analogs have shown a better PPGE control. However, there remains an unmet need with insulin analogs for a faster onset and a shorter duration of action, which could potentially help to achieve a better PPG control than the rapid-acting insulin analogs (11,12). Ultrafast-acting oral insulins can potentially simulate the first-phase insulin release after a meal and may lead to a better control of PPG and PPGE. Hence, ultrafast-acting oral insulins can help to correct the postprandial insulin deficiency in patients with type 2 diabetes (13).

Insulin Tregopil (hereafter referred to as Tregopil), is an ultrafast-onset, short-acting oral prandial insulin analog. It is a recombinant human insulin that contains a single, short-chain amphiphilic oligomer modified with the covalent attachment of methoxy-triethylene-glycol-propionyl moiety at Lys-β29-amino group of the B chain via an amide linkage (14). Tregopil restores insulin availability during the immediate post-meal period with a median time-to-peak pharmacodynamic effect, i.e., best glucose reduction being ∼30-40 min post-dose (14). It possibly mimics the release of insulin during the first-phase period and reduces early postprandial hyperglycemia (PPH) in patients with type 2 diabetes (14). Moreover, the efficacy of Tregopil on the initial PPG control has been well established in a sequential single ascending dose study versus placebo (14) and in a phase-3 randomized, placebo-controlled study in patients with type 2 diabetes on optimal doses of metformin (data on file). The current study evaluated the efficacy and safety of two doses of Tregopil (30 mg and 45 mg) compared to insulin Aspart (IAsp) in patients with type 2 diabetes being treated with stable doses of insulin glargine and metformin (+/- OADs).

## RESEARCH DESIGN AND METHODS

### Study Setting and Population

This study was a 24-week, open-label, randomized, parallel-group, phase 2/3 study (CTRI No.: CTRI/2017/11/010560; Clinicaltrials.gov identifier: NCT03430856) conducted across 20 centers in India between 2017 and 2019. Patients with an established diagnosis of type 2 diabetes (American Diabetes Association [ADA] 2017 guidelines; glycated hemoglobin [HbA_1c_] ≥ 6.5%) (15) for at least 6 months before screening and with HbA_1c_ levels between 7.5% and 10% (58-86 mmol/mol) at screening, and on a stable dose of metformin ± OADs ± basal insulin, and with HbA_1c_ levels between 7% and 9% (53-75 mmol/mol) at randomization, and aged between 18 and 70 years were enrolled into the run-in period (detailed inclusion and exclusion criteria are given in Supplementary Methods: A). All study participants provided written informed consent confirming their voluntary participation. The trial was approved by respective independent ethics committees/institutional review boards (details given in Supplementary Methods: B) and conducted in accordance with the Declaration of Helsinki, Indian Council of Medical Research Ethical guidelines for Biomedical Research, and Good Clinical Practices.

### Study Procedure

The trial duration was approximately 37 weeks (3 weeks screening period, 8 weeks run-in period, 24 weeks treatment, and 2 weeks safety follow-up).

### Randomization

At the end of the run-in period, patients meeting the randomization criteria (details specified in Supplementary Methods: C1) were randomized (1:1:1) to Tregopil (30 mg [*n* = 30], 45 mg [*n* = 31]) and IAsp (*n* = 30) in addition to optimized doses of insulin glargine and metformin (+/-OADs) (Supplementary Fig. S1). The randomization scheme was produced using a validated system and an interactive voice response system was used to assign treatment at each site. While the treatment allotment was open label, post baseline HbA_1c_ assessments were blinded to the investigators throughout the treatment period.

### Dosing and Dose Titration

Tregopil doses, 30 mg (2 tablets of 15 mg each) and 45 mg (3 tablets of 15 mg each), were administered orally thrice daily (TID), 10 ± 2 min before each of the three major meals in a day. SC IAsp injection (100 U/mL) was administered TID, within 5 min before each of the three major meals. Additional details on dosing are provided in Supplementary Fig. S2. During the first 4 weeks of the treatment period (active titration period), study doses were up-/down-titrated by the investigator as per prespecified titration algorithms based on self-monitored blood glucose (SMBG) levels and hypoglycemia criteria to achieve target PPG levels. The patients were instructed on the identified optimum dosage regimen and advised to follow it for the remainder of the treatment period. Additional information is provided in Supplementary Methods: C2.

### Study Endpoints

PPG-related efficacy assessments included PPG parameters (PPG and PPGE) assessed following standardized test meal ([STM] administered at breakfast, at baseline [randomization visit], and weeks 12, 20, and 24), 9-point SMBG (performed at baseline and weeks 8, 12, 16, and 24) and formed important secondary endpoints of the study. Fasting plasma glucose (FPG) was also assessed.

The primary endpoint was change from baseline (CFB) in HbA_1c_ levels at week 24. Secondary endpoints included the number of severe or clinically significant hypoglycemic events during the treatment period, classified according to ADA 2017 and Food and Drug Administration guidelines (15). Treatment-emergent adverse events (TEAEs) were summarized based on the severity of adverse events (AEs) and relationship to trial medication (16).

### Post-hoc analyses to evaluate different groups

Analyses were carried out to evaluate the relationship between patterns in plasma glucose response and HbA_1c_ as well as variations in PPG and HbA1c. Estimated HbA_1c_ (eA_1c_) was calculated using the formula: mean glucose (mg/dL) * 0.0348 + 1.6 (17) (details are provided in Supplementary Methods: C3). Additionally, based on 9-point SMBG levels at weeks 24 and 12, PPG parameters were analyzed using General Linear Mixed Models (PROC MIXED & PROC MEANS) along with corresponding two-sided *P*-value, 95% confidence interval (CI), and descriptive statistics.

### Safety

Safety evaluation included assessment of AEs and serious AEs (SAEs), symptomatic hypoglycemia, SMBG, 3-point SMBG, vital signs, and standard clinical laboratory evaluations (additional information is provided in Supplementary Methods: C4).

### Statistical Analysis

All randomized patients were included in the intent-to-treat analysis set (ITT), while the safety analysis set included randomized patients on at least one dose of the study drug. Per-protocol analysis set included all ITT patients who met all inclusion requirements, did not meet any exclusion criteria, had a baseline HbA_1c_ measurement, were exposed to the study medication, had at least one HbA_1c_ measurement at or after 8 weeks of randomization, and had no protocol deviation that affected the primary outcome. Patient follow-ups were carried out at study-specific timepoints for efficacy and safety assessments.

As this was a proof-of-concept exploratory study, no statistical rationale was defined for the sample size estimation. A minimum of 30 patients in each treatment group was deemed adequate to evaluate the study endpoints. Continuous variables were summarized using descriptive statistics as mean, median, standard deviation, and 95% CI (minimum and maximum). Categorical variables were summarized as counts and percentages. All statistical hypothesis tests were performed at a 5% level of significance (two-sided test). *P*-values <0.05 were considered statistically significant. All statistical analyses were performed using SAS^®^ version 9.4 (or higher) for Windows (SAS Institute Inc., Cary, North Carolina, USA).

## RESULTS

Overall, 91 patients were randomized to the Tregopil (30 mg [*n* = 30], 45 mg [*n* = 31]) and IAsp groups (*n* = 30), of which 85 patients completed the trial (Supplementary Fig. S1). Baseline and clinical characteristics were balanced between the treatment groups (Supplementary Table S1).

### Postprandial Glucose Control

#### Standardized Test Meal

##### PPG

Baseline mean PPG values up to 1-h and up to 0-4 h post meal for all the treatment groups are shown in Fig. 1a and 1b, respectively and the mean data for the same at week 24 are given in Fig. 1c and 1d. Similarly, the mean and CFB of PPG levels from 1-h to 4-h at week 24 are summarized in Table 1 (the 1-h and 2-h estimated treatment differences [ETD] for PPG are provided in Supplementary Table S2). At week 24, the mean 1-h PPG levels were numerically lower and the corresponding CFB greater with Tregopil, indicating more effective control compared to IAsp (Table 1 and Fig. 1c), while the mean 2-h PPG and CFB levels were comparable (Table 1 and Fig. 1d), indicating control as effective as IAsp. The mean week 24 3-h and 4-h PPG were quantitatively lower and the corresponding CFB was greater in the IAsp group versus the Tregopil groups (Fig. 1d). A similar trend in the mean PPG and CFB levels was observed at week 12; however, no statistically significant differences in any of the parameters were observed between the treatment groups. Although glucose control in the IAsp group was numerically better in the later post meal period, i.e., at 3-h and 4-h, area under the curve (AUCs) for glucose over the 4-h post meal period of the Tregopil and IAsp groups were similar (Table 2). Tregopil groups had a higher proportion of patients meeting the more stringent 2-h PPG target level (<140 mg/dL) at week 24 (30 mg, 21.4%; 45 mg, 26.7%) compared to the IAsp group (17.2%) (Supplementary Fig. S3). The overall PPG improvement occurred despite a slight worsening of FPG from baseline in the Tregopil groups vs similar levels maintained or slight improvement in FPG in the IAsp group.

**Table 1:** Summary of postprandial plasma glucose (PPG) and postprandial plasma glucose excursions (PPGE) at different post mealtime points during STM at week 24

|  |  |  | Tregopil 30 mg<br>(N = 30) | Tregopil 45<br>mg<br>(N = 31) | IAsp<br>(N = 30) | Tregopil 30 mg<br>+ 45 mg<br>(N = 61) |
| --- | --- | --- | --- | --- | --- | --- |
| PPG (mg/dL) |  |  |  |  |  |  |
| Week 24 | 0 min | Mean | 131.20 (43.42) | 118.60 (27.10) | 114.90 (26.20) | - |
|  |  | CFB | 27.00 (37.76) | 9.80 (41.01) | -6.50 (35.86) | - |
|  | 1-h | Mean | 168.50 (69.34) | 157.90 (52.88) | 170.10 (41.74) | - |
|  |  | CFB | -34.40 (67.52) | -29.90 (54.86) | -25.00 (50.76) | - |
|  | 1.5-h | Mean | 184.70 (75.76) | 173.60 (53.69) | 181.20 (44.49) | - |
|  |  | CFB | -25.90 (82.19) | -27.40 (58.54) | -29.80 (57.42) | - |
|  | 2-h | Mean | 187.70 (74.54) | 185.50 (60.85) | 180.40 (45.98) | - |
|  |  | CFB | -29.30 (94.67) | -23.40 (60.60) | -34.20 (60.79) | - |
|  | 3-h | Mean | 173.10 (54.87) | 172.20 (50.67) | 158.10 (50.01) | - |
|  |  | CFB | -19.00 (90.24) | -20.30 (56.20) | -42.40 (55.54) | - |
|  | 4-h | Mean | 150.70 (50.29) | 161.40 (61.04) | 136.30 (52.06) | - |
|  |  | CFB | -20.20 (84.25) | -3.80 (57.30) | -32.20 (49.78) | - |
|  |  |  | Mean (SD)<br>n = 27 | Mean (SD)<br>n = 30<sup>#</sup> | Mean (SD)<br>n = 27* | Mean (SD)<br>n = 57 |
| PPGE (mg/dL) |  |  |  |  |  |  |
| Week 24 | 1-h | Mean | 37.30 (50.30) | 39.37 (47.69) | 55.15 (34.56) | 38.39 (48.51) |
|  |  | CFB | -61.44 (45.84) | -39.73 (55.97) | -16.11 (37.46) | -50.02 (52.13) |
|  | 1.5-h | Mean | 53.56 (55.13) | 55.07 (45.13) | 66.30 (36.13) | 54.35 (49.66) |
|  |  | CFB | -52.89 (59.93) | -37.23 (61.73) | -21.63 (37.88) | -44.65 (60.86) |
|  | 2-h | Mean | 56.56 (54.41) | 66.90 (53.27) | 65.52 (34.58) | 62.00 (53.58) |
|  |  | CFB | -56.30 (77.51) | -33.20 (67.09) | -26.07 (42.50) | -44.14 (72.46) |
|  | 3-h | Mean | 41.89 (43.30) | 53.60 (54.57) | 43.42 (41.79) | 48.05 (49.47) |
|  |  | CFB | -46.07 (78.74) | -30.10 (71.03) | -37.27 (43.91) | -37.67 (74.54) |
|  | 4-h | Mean | 19.56 (39.49) | 42.87 (56.46) | 21.62 (45.15) | 31.82 (50.13) |
|  |  | CFB | -47.22 (72.64) | -13.67 (59.37) | -28.50 (44.39) | -29.56 (67.53) |

\*n = 26 for 3-h and 4-h actual PPGE and CFB in IAsp group; \* n = 26 for 3-h and 4-h actual PPG in IAsp group; n = 25 for 3-h and 4-h CFB in IAsp group.
Baseline was defined as the last observed value of a parameter before first intake of trial medication. Change from baseline (CFB) was calculated as the difference between value of interest and corresponding baseline value.
PPGE - Post prandial glucose excursion; STM - Standardized test meal; CFB - Change from baseline STM; SD - Standard deviation; IAsp - Insulin Aspart.

**Table 2:** Mean differences and T/R ratio (%) of AUC for Tregopil 30 mg + 45 mg vs IAsp following a STM at week 24

| Timepoints at week 24 | LSMD [95% CI] |  |  | T/R ratio (%) |  |
| --- | --- | --- | --- | --- | --- |
|  | Tregopil 30 mg vs IAsp | Tregopil 45 mg vs IAsp | Tregopil 30 mg + 45 mg vs IAsp | Tregopil 30 mg Vs IAsp | Tregopil 45 mg vs IAsp |
| **AUC (0-1h)** | -1109<br>[-2148, -68.75] | -702.7<br>[-1743, 337.15] | -905.7<br>[-1812, 1.12] | 96.47 | 95.95 |
| **P value** | 0.037 | 0.183 | 0.050 |  |  |
| **AUC (0-2h)** | -2070<br>[-4505, 364.96] | -1146<br>[-3581, 1289.3] | -1608<br>[-3731, 515.46] | 100.20 | 97.30 |
| **P value** | 0.095 | 0.353 | 0.136 |  |  |
| **AUC (0-3h)** | -2540<br>[-6356, 1275.2] | -628.4<br>[-4444, 3187.1] | -1526<br>[-4428, 1376.6] | - | - |
| **P value** | 0.190 | 0.745 | 0.348 |  |  |
| **AUC (0-4h)** | -2487<br>[-7519, 2544.7] | 473.77<br>[-4558, 5505.4] | -1007<br>[-5394, 3381.1] | 104.29 | 103.20 |
| **P value** | 0.330 | 0.852 | 0.650 |  |  |

IAsp- Insulin Aspart, AUC - Area under curve; STM – Standardized test meal; LSMD - Least squared mean difference; 95% CI - 95% confidence interval

**Figure 1:**
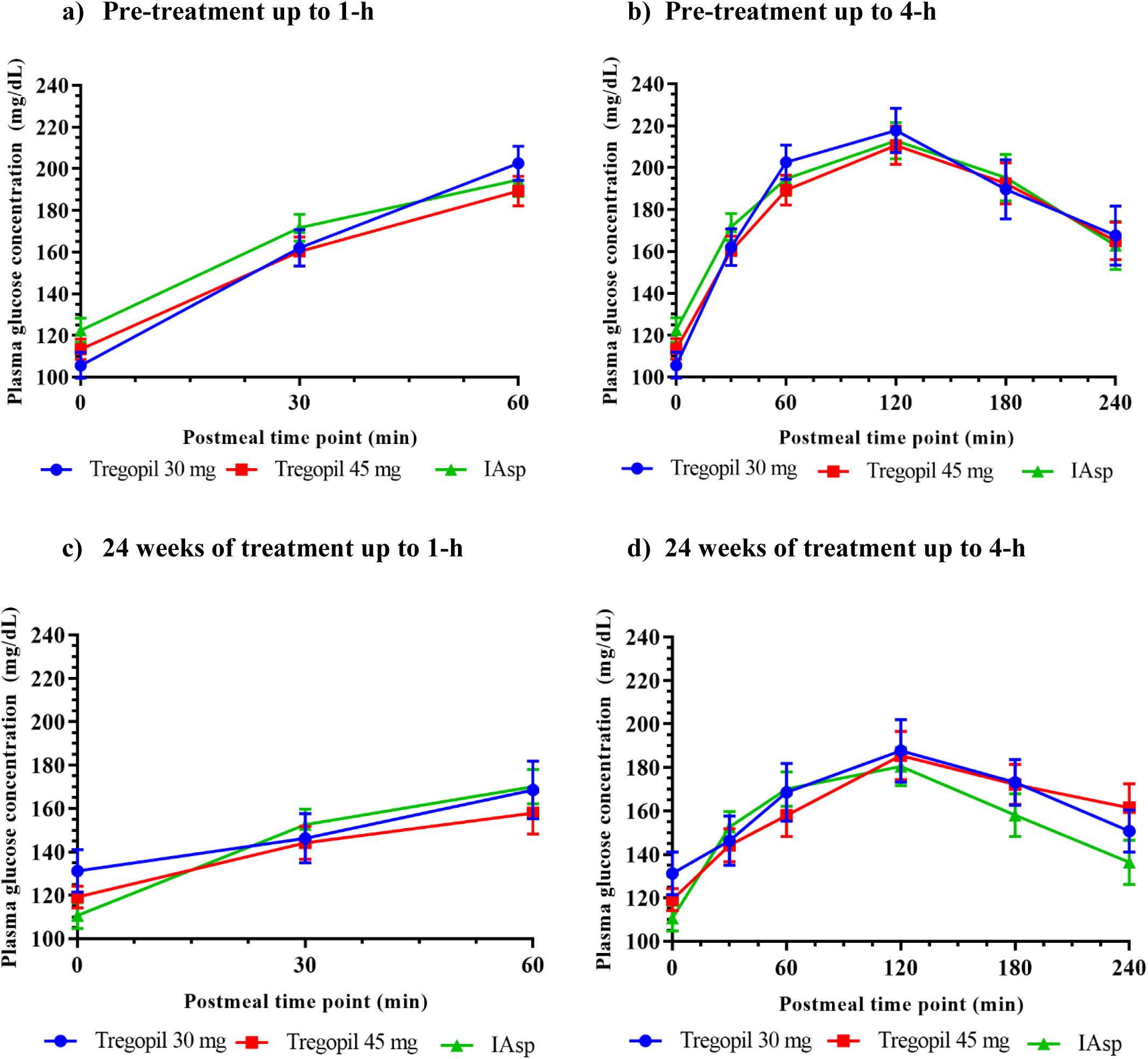
STM - Mean plasma glucose concentration (PPG) vs time - a) at pre-treatment up to 1-h; b) at pre-treatment up to 4-h; c) at week 24 up to 1-h; d) at week 24 up to 4-h Mean with SEM for PPG is presented in the above graphs. Post meal timepoint - 0 min indicates, 10 minutes after the dose, i.e., meal start time.

##### PPGE

At week 24, 1-h PPGE (mean and CFB) showed a trend towards more effective control in Tregopil groups compared to IAsp group (Table 1), with the Tregopil 30 mg group (ETD) [95% CI], -45.33 mg/dL (−71.91, -18.75), *P* = 0.001) and the combined Tregopil group (30 mg + 45 mg) (−33.91 mg/dL [-56.72, -11.09], *P* = 0.004) showing a significant difference (Supplementary Table S2). The week 24 mean PPGE levels at 1.5-h and 2-h in Tregopil groups were controlled as effectively as in the IAsp group (Table 1). However, Tregopil groups displayed a trend towards better reduction of 1-h, 1.5-h, and 2-h PPGE from baseline compared to IAsp. At 3- and 4-h, the PPGE levels (mean and CFB) were similar in the combined Tregopil group and the IAsp group. At week 12, similar trends were observed except at 1.5-h, where IAsp showed better reduction than Tregopil (30 mg).

Additional analyses showed that Tregopil (combined dose group) was associated with lower PPGE at 15 min compared to IAsp; however, the difference was not statistically significant (week 24, 9.46 mg/dL vs. 21.46 mg/dL; *P* = 0.076). The lower mean PPGE in Tregopil groups (combined) continued through 30- and 45-min post meal at week 24 for both combined Tregopil and IAsp groups (30 min, 20.65 mg/dL vs. 39.70 mg/dL; *P* = 0.022; 45 min, 29.00 mg/dL vs. 45.44 mg/dL; *P* = 0.109), thereby demonstrating the faster onset of action of Tregopil. Similar differences in the mean PPGE were observed at week 12 between combined Tregopil and IAsp groups (30 min, 10.9 mg/dL vs. 33.0 mg/dL; *P* = 0.002; 45 min, 20.0 mg/dL vs. 40.8 mg/dL; *P* = 0.023).

Post meal glucose AUC (by STM) at week 24 was lower in the combined Tregopil group compared to the IAsp group at 1-h (AUC_0-1h_, *P* = 0.050) and, at week 12, similar significant reductions in AUC were observed at 1-h and 2-h (AUC_0-1h_, *P* = 0.004; AUC_0-2h_, *P* = 0.033). Also, AUC_0-3h_ and AUC_0-4h_ were similar between the treatment groups at week 24 (Table 2).

#### 9-Point Self-Monitoring Blood Glucose

##### PPG

At week 24, mean PPG levels were comparable between the Tregopil and IAsp groups at 1 and 2 h post breakfast and 1 h post lunch (Supplementary Table S3). PPG levels were significantly higher in the Tregopil groups than in the IAsp group at 2 h post lunch and 1 and 2 h post dinner. However, the CFB in mean PPG at different post meal timepoints (Supplementary Fig. S4) were comparable between treatment groups except for the 2 h post dinner value favoring IAsp versus both the Tregopil groups, 30 mg (*P* = 0.020) and 45 mg (*P* = 0.047) (Supplementary Table S4).

The proportion of patients who achieved 2-h PPG <140 mg/dL at week 24 (from SMBG) was numerically higher in the Tregopil groups (30 mg, 25.0%: 45 mg, 23.3%) than the IAsp group (17.2%). Similarly, the proportion of patients who achieved 2-h PPG <140 mg/dL following STM was numerically higher in the Tregopil groups (30 mg, 21.4%; 45 mg, 26.7%) compared to the IAsp group (17.2%) (Supplementary Fig. S3). At the end of the titration period, despite the highest allowable dose (45 mg TID) in the study, the Tregopil 45 mg group had >60% of patients (with 2-h PPG by SMBG >180 mg/dL) indicating the need for a higher dose.

##### PPGE

Overall, at week 24, the 1-h and 2-h PPGE (mean and CFB) at breakfast, lunch, and dinner did not show statistically significant differences between the treatment groups. The CFB in all 1-h post meal PPGE and 2-h post breakfast excursion showed a trend towards more effective control in the Tregopil groups than IAsp (Fig. 2 a, b, and c). PPGE was numerically lower in the Tregopil groups at 2-h post breakfast, but CFB was better for IAsp. The mean 1- and 2-h PPGE at week 24 is represented in Supplementary Table S3.

**Figure. 2:**
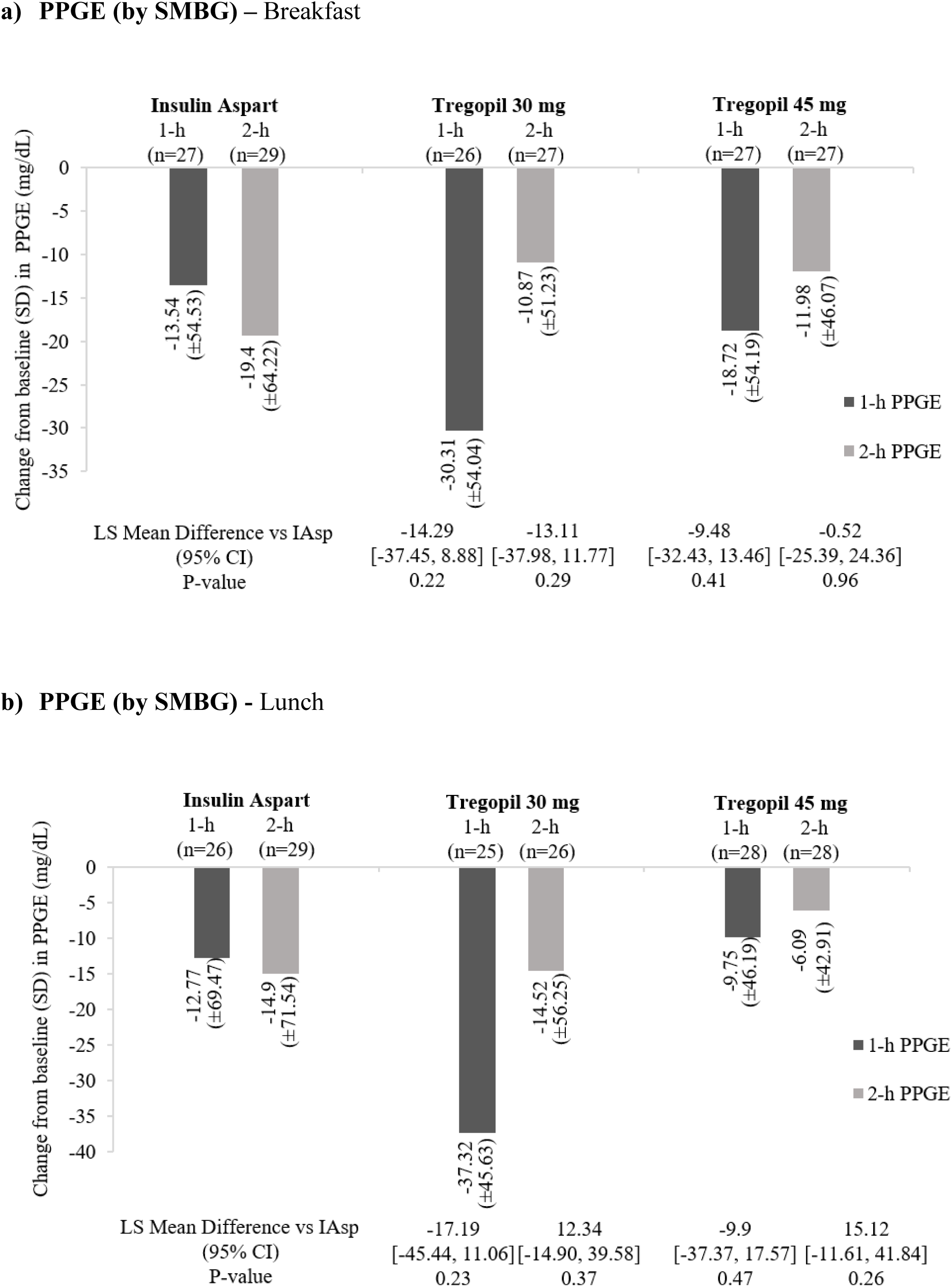
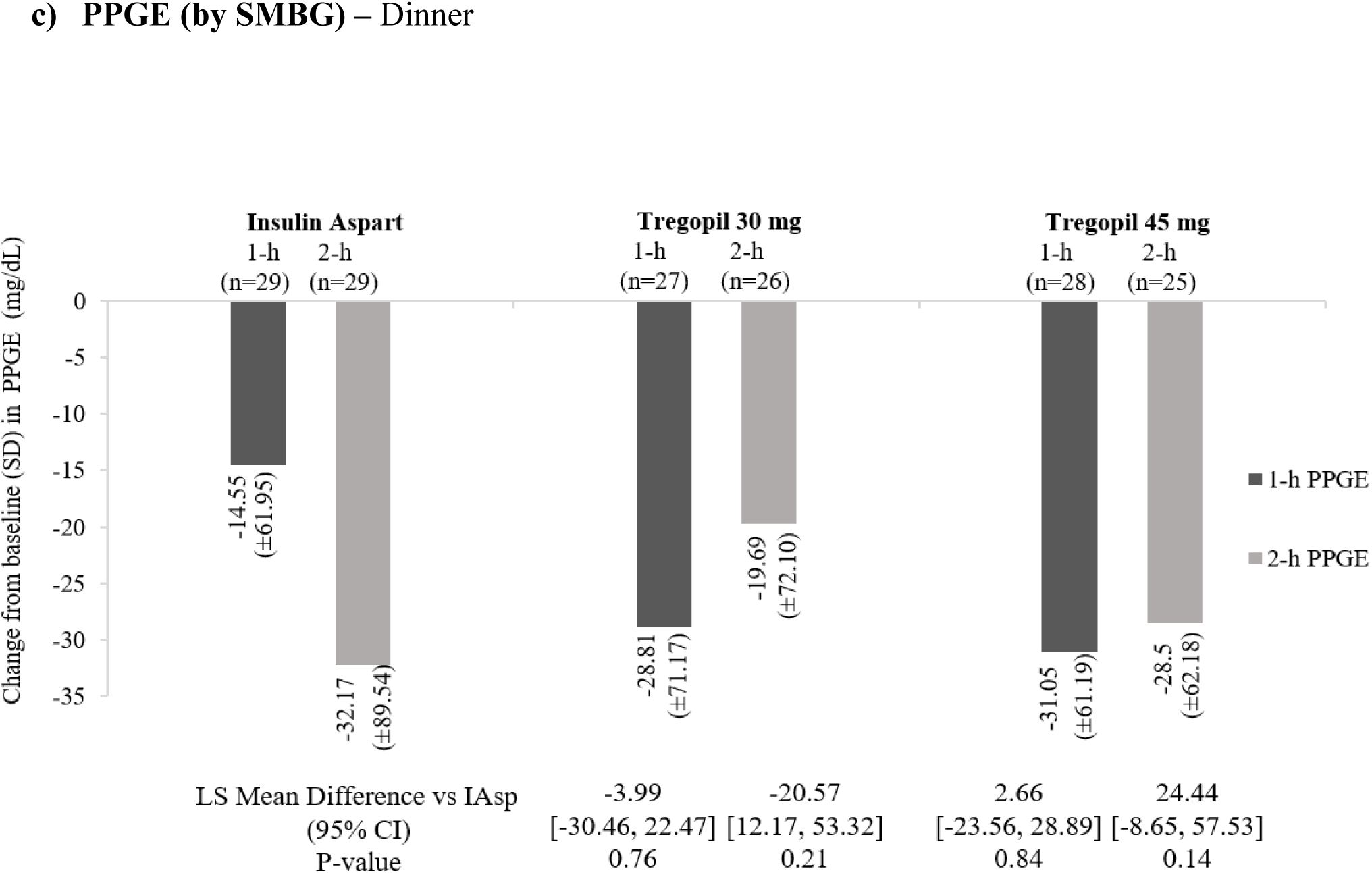
CFB in PPGE levels (1 and 2 h) from 9-point SMBG at week 24 following each meal of the day. a) CFB in PPGE by SMBG at breakfast; b) CFB in PPGE by SMBG at lunch; c) CFB in PPGE by SMBG at dinner. Baseline was defined as the last observed value of a parameter before first intake of trial medication. Change from baseline (CFB) was calculated as the difference between value of interest and corresponding baseline value.

### HbA_1c_

The mean CFB in measured HbA_1c_ at weeks 24 and 12 for the Tregopil groups was 0.15% and -0.07% for 30 mg and 0.22% and -0.17% for 45 mg, respectively. For the IAsp group, the mean CFB for measured HbA_1c_ at weeks 24 and 12 were -0.77% and -0.88%, respectively.

The HbA_1c_ analysis, interestingly revealed a discordance in the mean glucose-HbA1c in the Tregopil groups but not in the IAsp group. To further explore this finding, the measured HbA_1c_ was compared to estimated A1c (eA_1c_) calculated from the mean overall daily blood glucose measurements based on 9-point SMBG using previously published methodologies (17). The results showed a reduction from baseline in eA1c at week 24 in the insulin Tregopil groups as opposed to the slight increase seen with the central laboratory measured HbA1c. However, this discrepancy between eA1c and measured A1c was not observed in the IAsp group (Supplementary Table S5).

### Changes in Fasting Plasma Glucose and Body Weight

FPG increased from baseline in Tregopil groups but not in the IAsp group. At week 24, the mean FPG increased from 107.2 ± 31.0 to 132 ± 5 mg/dL (CFB: +24.1 mg/dL) in the Tregopil 30 mg group and from 109.7 ± 27.3 to 120 ± 27 mg/dL (CFB: +11.8 mg/dL) in Tregopil 45 mg group, while the FPG decreased from 120.5 ± 32.2 to 113 ± 29 mg/dL (CFB:-9.6 mg/dL) with IAsp.

### Effect on body weight

There was no significant change in mean body weight in any of the treatment groups during the treatment period in this study.

### Safety

#### Adverse events

Overall, Tregopil was safe and well tolerated. The proportion of patients with at least one TEAE in the Tregopil groups (30 and 45 mg) were 20.0% and 16.1% respectively and with IAsp, 13.3% (Supplementary Table S6). Of the total population, 16.5% patients reported overall 29 TEAEs. Majority of the TEAEs (21 of 29; most common: pyrexia and gastritis) were mild in intensity (Tregopil 30 mg [*n* = 4] 4 events; Tregopil 45 mg [*n* = 4] 10 events; IAsp [*n* = 4] 7 events). Seven moderate-intensity TEAEs were reported (Tregopil 30 mg [*n* = 2] 3 events, Tregopil 45 mg [*n* = 1] 4 events). No severe TEAEs or SAEs were reported in the study.

#### Hypoglycemia

The percentage of patients who reported any hypoglycemic events across Tregopil groups (30 and 45 mg) were 86.7% and 83.9%, respectively, and 83.3% with IAsp. The rate (events/per 100 years of exposure) of clinically significant (level 2: ADA Hypoglycemic Episode Level 2 Definition: Glycemic level < 54 mg/dL; Sufficiently low to indicate serious, clinically important hypoglycemia) treatment-emergent hypoglycemia events in the Tregopil groups (30 mg, 282.9; 45 mg, 193.3) were lower compared to the IAsp group (346.3) (combined Tregopil groups vs. IAsp, rate ratio [RR]: 0.69). The incidence of hypoglycemic events during each post meal period was lower in the Tregopil groups than with IAsp (Supplementary Table S6).

## CONCLUSIONS

Tregopil is an ultrafast, short-acting, oral prandial insulin with a peak action at around 30-40 min and a duration of action of 2-3 h. Tregopil resulted in an improved control of PPH over a 4-h post-meal period in patients with type 2 diabetes when administered 10 min before major meals, in addition to optimized and stable basal insulin and other medications.

In this study, Tregopil at a 45 mg daily dose resulted in an average 1-h PPG level of 157.9 mg/dL, which was close to the required 1-h cut-off value of 155 mg/dL (18). Uncontrolled PPG is considered an important predictor of developing diabetes and is linked to inflammation, thrombosis, endothelial dysfunction, and oxidative stress generation, all of which may contribute to the pathogenesis of cardiovascular disease (19,20).

The STM assessment (at breakfast) showed that the oral Tregopil reduced 1-h and 2-h PPG levels and excursions as effectively or, in some instances (early post meal period), more effectively than premeal IAsp. As expected, Tregopil’ s effect on 3-h and 4-h PPG reduction was less than IAsp. However, cumulative glucose control over the 4-h post meal period (AUC) was comparable between Tregopil and IAsp.

Similarly, results of the 9-point SMBG studies at baseline and at 12 and 24 weeks of treatment showed Tregopil to be (1) as effective or more effective compared to IAsp in reducing PPG levels and excursions 1-h and 2-h after breakfast; (2) as effective in lowering 1-h PPG and excursions at lunch but (3) trending towards less effective control of 2-h PPG after lunch and 1-h and 2-h PPG after dinner. The effect of Tregopil was greatest during breakfast, lower at lunch, and least during the evening meal. The study’s most significant finding is that Tregopil works particularly well at breakfast, a meal with a high physiological requirement of insulin. Higher Tregopil doses or the addition of supplementary basal insulin to regulate basal glucose levels (i.e., fasting and between-meals) can compensate for the suboptimal control during later meals of the day, resulting in better glycemic control in patients with type 2 diabetes on the Tregopil regimen.

The results indicate that Tregopil could provide a glycemic control comparable to IAsp in patients with type 2 diabetes who have significantly elevated PPG despite near-normal FPG (<120 mg/dL based on SMBG). A post-hoc responder analysis for different % cut offs of HbA1c at week 24 showed: 1) A clinically relevant 0.3% reduction in HbA1c from baseline in 40% patients in the Tregopil 30 mg group and 45.1% in 45 mg group versus 66.6% in the IAsp group. 2) Any reduction in HbA1c in nearly 50% of patients in the Tregopil 30 mg group, 45.6% in 45 mg group and 86.7% in IAsp group. A further subgroup analysis demonstrated that decrease in the measured HbA_1c_ from baseline was greatest in patients with well-controlled FPGs, emphasizing the importance of FPG in reflecting the effect of PPG control on HbA_1c_ reduction (14). This is especially true with Tregopil, which, unlike IAsp, has a shorter duration of glucose-lowering activity. Tregopil’s shorter time action profile also explains the modest increase in FPG associated with Tregopil but not with IAsp therapy throughout the 24-week treatment. The effect of IAsp begins 15 minutes after the injection and lasts for 4-6 hours. IAsp, due to its longer duration of action, reduces inter-meal glucose levels to some extent, lessening the need for basal insulin, while also predisposing the patient to late-phase hypoglycemia. Adding Tregopil to a regimen of basal insulin and OADs with continued FPG optimization may result in a better overall glycemic control and not just PPG improvement as observed in this study. Oral insulin Tregopil can be especially useful in type 2 diabetes patients on basal+, basal++ or bolus regimens in subgroup of patients with good FPG control to further improve overall glycemic control.

Tregopil may be potentially beneficial in special settings such as when patients need insulin for PPG control during travel or other short-term situations where use of injectable formulations is inconvenient.

It is well known that HbA_1c_ measurements can be influenced by factors (21) other than glucose levels, such as hemoglobinopathies, red blood cell (RBC) survival (22), and metabolic factors influencing the glycation reaction. Information about glycemic variability or differentiation among fasting, pre-prandial and post-prandial glycemia is not accurately reflected by HbA_1c_ (23). It is not surprising, therefore, that HbA_1c_ may not accurately represent the improvement in overall daily mean blood glucose levels, especially PPH reduction in Tregopil-treated patients. In IAsp-treated patients, the improvement in glucose control was reflected by an appropriate reduction in HbA_1c_ measurement. This difference indicates that HbA_1c_ may not be a complete measure of glycemic control in patients treated with Tregopil. Our observations that plasma glucose variations over time may not be fully reflected in HbA_1c_ measurements underscores the need of exploring alternative glycemic markers.

Oral insulin absorbed from the gastrointestinal tract with an ultrafast onset-of-action could effectively target excessive and prolonged PPG excursions and can limit insulin exposure to peripheral tissues, thereby reducing hypoglycemia risk and weight gain. The duration and size of this study limited the evaluation of Tregopil’s effect on body weight. However, it shows a potential towards a lower risk of hypoglycemia associated with Tregopil treatment than an injected prandial insulin.

Strengths of the present study include a run-in period resulting in optimized basal insulin and antidiabetic medications and a more homogeneous population at randomization; an active-controlled study with prandial IAsp with appropriate titration; stable maintenance period to evaluate sustenance of use; and application of both standardized meal test and real-life scenario for evaluation (SMBG) of PPG control. This was a well-monitored study with standard of care counseling, including lifestyle modifications. A titration review committee monitored the dose titrations throughout the study.

The major limitation of the study is its open-label design due to difference in routes of administration of the test and control drugs and the ethical challenges of the double-dummy design due to the daily administration of three SC injections of active control drug. The requirement of up-titration allowed up to a maximum of 8 weeks to achieve desired glycemic control was high, as ∼90% of patients in the Tregopil 30 mg group were up-titrated to 45 mg for at least one of the doses daily. Additionally, the small size of the study population limited the statistical power and interpretation of results.

In conclusion, this study demonstrates the efficacy of oral Tregopil in controlling PPG excursions with a lower incidence of clinically significant hypoglycemia compared to IAsp in patients with type 2 diabetes on a basal-bolus regimen. Tregopil had a more rapid onset and a shorter duration of action than IAsp. Tregopil is effective at controlling PPG excursions; however, further modifications in basal insulin dosage may be required for good FPG management. These features, along with early PPH control and low hypoglycemia risk, make oral Tregopil a potentially attractive treatment option for type 2 diabetes, especially in patients with a good FPG control.

## Data Availability

All data produced in the present work are contained in the manuscript

## Acknowledgments

The authors thank all investigators and study participants for generously contributing their time for this study. The authors acknowledge support by Hema Balasubramanian (Biocon Biologics Ltd) and Vasan Sambandamurthy and Nikhil Dixit (ex-employees of Biocon) for their contribution towards clinical operation activities related to this study. The authors extend their thanks to Vathsala Jayanth, MD (Biocon Biologics Ltd) for medical writing support under their guidance. The authors acknowledge Molecular Connection Pvt. Ltd., Bengaluru, India for editorial services towards development of this article.

## Author Contributions

H.E.L., A.F., A.D.C., S.N.A., A.V., S.L., and S.R.J were involved in study conceptualization and design. A.M. was involved in data curation and data validation. J.P. was involved in project administration. All authors analyzed and interpreted the study data and results. All authors participated in the preparation and review of the manuscript. All authors read and approved the final version of the manuscript.

