## Supplementary Material for "Efficacy and Safety of Tregopil, a Novel, Ultra-Rapid Acting Oral Prandial Insulin Analog, as Part of a Basal-Bolus Regimen in Type 2 Diabetes: A Randomized, Active Controlled Phase 3 Study"

**Supplementary Data**

**Supplementary A:** Study inclusion and exclusion criteria

**Supplementary B:** Details of respective independent ethics committees/institutional review boards of the study

**Supplementary C:** Methods

C1. Randomization criteria

C2. Dosing and dose titration

C3. Post-hoc analyses of HbA_1c_

C4: Additional study endpoints

**Supplementary Table S1:** Demographic and clinical characteristics of study participants at baseline

**Supplementary Table S2:** Analyses of 1-h and 2-h Postprandial Plasma Glucose Excursion (PPGE) and Postprandial Plasma Glucose (PPG) by Standard Meal Tolerance (STM) at week 24

**Supplementary Table S3:** Summary of 1-h and 2-h PPGE and PPG from 9-point SMBG at week 24

**Supplementary Table S4:** Analyses of 1-h and 2-h PPGE and PPG by SMBG at week 24

**Supplementary Table S5:** Mean measured HbA_1c_ and estimated A_1c_ (eA1c) at baseline and at week 24

**Supplementary Table S6:** Incidences of hypoglycemic events and TEAEs during the 24-week study period

**Supplementary Figure S1:** Patient disposition

**Supplementary Figure S2:** Study design

**Supplementary Figure S3:** Percentage of patients achieving 2-h PPG level (<140 mg/dL) after STM and/or measured by 9-point SMBG at week 24

**Supplementary Figure S4:** CFB in PPG levels (1-h and 2-h) from 9-point SMBG at week 24 following each meal of the day

**Supplementary A:** Study inclusion and exclusion criteria

**Inclusion criteria**

1. Male and female patients between the ages of 18 to 70 years, who had provided written informed consent for participation in the trial and were willing to comply with trial procedures.
2. Patients with an established diagnosis of type 2 diabetes mellitus and a duration of diabetes mellitus of at least 6 months at screening based on criteria given below as per American Diabetes Association (ADA) 2017 guidelines:

- HbA_1c_ ≥ 6.5% (OR)
- FPG ≥ 126 mg/dL (OR)
- 2-hrs PPG level of ≥ 200 mg/dL during an oral glucose tolerance test. The test was to be performed as described by the World Health Organization, using a glucose load containing the equivalent of 75 g anhydrous glucose dissolved in water (OR)
- In a patient with classic symptoms of hyperglycemia or hyperglycemic crisis, a random plasma glucose ≥ 200 mg/dL.

(If there was a suspicion, the investigator did a fasting plasma C-peptide [Levels < 0.3 nmol/L was indicative of Type 1 Diabetes Mellitus and the patient was excluded]).

3. Patients should have been on a stable dose of metformin (at least 1500 mg daily [daily dose of at least 1000 mg was permitted if intolerant to 1500 mg dose]) for a period of at least 3 months prior to screening.
4. Patients who were eligible for initiation of or were already receiving insulin glargine.
5. Body mass index of 18.5 to 35.0 kg/m^2^.
6. Patients on stable diet and physical activity practices in the 3 months prior to screening with stable weight, (with no more than 5 kg gain or loss), in the 3 months prior to screening; this information was obtained from the patient history.
7. Hemoglobin ≥ 10.0 g/dL.
8. HbA_1c_ of 7.5 to 10.0%.
9. All women of childbearing potential (i.e., pre-menopausal) used at least 2 reliable forms of contraception during the trial, one of which must have been a physical barrier method and should have agreed to continue the contraceptive measures during the trial and for at least 10 days after receiving the last dose of the investigational medicinal product (IMP) /comparator drug:

- Periodic abstinence (e.g., calendar method, ovulation-symptothermal, and post-ovulation methods) and withdrawal method were not acceptable methods of contraception.
- Post-menopausal females must have had no regular menstrual bleeding for at least 1 year prior to screening. Follicle-stimulating hormone (FSH) test could have been done for confirmation if necessary, as per investigator discretion (Plasma FSH level of > 40 IU/L was indicative of menopause).
- Female patients who reported surgical sterilization must have undergone the procedure at least 6 months prior to screening. If not, appropriate contraceptives should have been used as described above.

10. All female patients of childbearing potential (irrespective of sterilization status) must have had negative serum pregnancy test result at screening.
11. Male patients must have been using 2 acceptable methods of contraception one of which must have been a physical barrier method, (e.g., spermicidal gel plus condom; condom plus partner was sterilized at least 6 months prior) for the entire trial duration and for at least 10 days following the last IMP/comparator drug administration.

**Exclusion criteria**

1. History or presence of a medical condition or disease that in the investigator’s opinion would have placed the patient at an unacceptable risk from study participation.
2. History of hypersensitivity to any of the active or inactive ingredients or excipients of Tregopil, insulin analog preparations (including IAsp), and metformin.
3. Patients known to be positive for autoimmune antibodies indicative of type 1 diabetes mellitus.
4. Treatment with glucagon-like peptide 1 agonists within 12 weeks before screening.
5. History of regular use (> 2 weeks of continuous therapy) of any premix or prandial insulin before the screening.
6. Ongoing treatment with OADs (e.g., thiazolidinedione's) contraindicated or unapproved for combination treatment with insulin (according to applicable product information) at screening visit.
7. Presence of gastrointestinal (GI) disorders or conditions known to significantly alter the absorption of orally administered drugs or significantly alter upper GI or pancreatic function including but not limited to GI motility disorders like gastroesophageal reflux disease, irritable bowel syndrome, inflammatory bowel disease (Crohn’s disease).
8. Patients who were likely to begin use of drugs including but not limited to colesevelam, metoclopramide, pramlintide, acarbose, domperidone; which might have interfered the absorption of oral drugs at the time of screening or within 5 times the half-life of the drug, whichever was longer.
9. History of ≥ 2 episodes of severe hypoglycemia (as per ADA 2017) within 6 months of screening and/or presence of hypoglycemia unawareness as judged by the investigator.
10. History of > 1 episode of hyperglycemic hyperosmolar coma or hospitalization for uncontrolled diabetes (e.g., diabetic ketoacidosis), within the 6 months prior to screening.
11. Any clinically significant abnormality in 12-lead electrocardiogram or safety laboratory tests deemed clinically relevant by the investigator at screening.
12. Serological evidence of human immunodeficiency virus, hepatitis B (HbsAg) or anti-hepatitis C antibodies at screening.
13. History of drug or alcohol dependence or abuse during the 1 year prior to screening.
14. Had received another investigational drug within 90 days (or as per local regulations, if any) prior to screening or if the screening visit was within 5 half-lives of another investigational drug (whichever was longer).
15. Previous participation in this study (participation was defined as enrolment in the run-in period).
16. Patients with the following secondary diabetic complications:

- Active proliferative retinopathy as confirmed by a dilated ophthalmoscopy (by the investigator, ophthalmologist or optometrist).
- Renal dysfunction indicated by modification of diet in renal disease, estimated glomerular filtration rate < 45 mL/min/1.73 m2 and/or diabetic nephropathy and/or clinical nephrotic syndrome at screening.
- History or existence of severe form of neuropathy or signs and symptoms of severe cardiac autonomic neuropathy.
- Patients with non-traumatic amputation (at any time) or clinically significant vascular procedure as a complication of diabetes within 1 year prior to screening.
- History of diabetic foot ulcer or non-healing diabetic ulcers in the 1 year prior to screening.

17. Any elective surgery requiring hospitalization planned during the study period.
18. Clinically significant major organ disease at the time of screening includes but not limited to:

- Uncontrolled (despite treatment) or untreated severe hypertension defined as systolic blood pressure above or equal to 180 mmHg or diastolic blood pressure above or equal to 100 mmHg (Stage 2 as per American Heart Association classification of hypertension)
- Uncontrolled hyperlipidemia (total cholesterol > 400mg/dL or very high serum triglycerides [≥ 500 mg/dL] levels as per American Association of Clinical Endocrinologists classification)
- Uncontrolled hyperthyroidism or hypothyroidism as per investigator’s assessment (investigator may conduct thyroid function test, if required for assessment).
- Impaired hepatic function (alanine aminotransferase or aspartate aminotransferase value > 2 times the upper limit of the reference range or serum bilirubin > 1.5 times the upper limit of the reference range).

19. Clinically significant cardiovascular and/or cerebrovascular disease within 12 months before screening including, but not limited to unstable angina, myocardial infarction, class III or class IV congestive heart failure according to the New York Heart Association criteria, valvular heart disease, cardiac arrhythmia requiring treatment, pulmonary hypertension, cardiac surgery, coronary angioplasty, stroke or transient ischemic attack.
20. Patients who taken in the last 12 months or may have required during the study period, systemic (including oral, intravenous, intramuscular) glucocorticoid therapy for more than 2 consecutive weeks.
21. History of cancer within the past 5 years prior to screening, except successfully treated non-melanoma skin cancer or cervical carcinoma in situ.
22. Patients who had donated blood or plasma within 12 weeks prior to screening (450 mL blood or equivalent).
23. Patients having hematological disorders (including but not limited to sickle cell disease, acquired and inherited hemolyticanemias, severe iron deficiency anemia) which could affect the HbA_1c_ assessment.
24. Patients who took acetaminophen containing medications on a regular basis and were unable or unwilling to substitute with a medication not containing acetaminophen the day before placement of the sensor and throughout each 7-day continuous glucose monitoring sensor periods.
25. Patients, who in the opinion of the investigator, may not have been able to understand and/or comply with the study procedures including answering the study questionnaires, maintaining a stable diet and exercise pattern (e.g., tendency to go on prolonged fast), frequent time zone travels etc., were excluded from the study.

**Supplementary Methods B:** Details of ethics committees of study sites have been listed below. The study has been approved by all ethic oversight bodies mentioned below:

1. Nirmal Hospital Pvt. Ltd. Ethics Committee, Ring Road, Surat-395 002, India
2. Institutional review Board (IRB), Christian Medical College, Vellore, India
3. Institutional Ethics Committee, Eternal Heart Care Center & Research Institute, Jaipur, Rajasthan, India
4. Institutional Ethics Committee, Madras Diabetes Research Foundation, 4, Conron Smith Rd, Gopalapuram, Chennai, Tamil Nadu 600086, India
5. Medilink Ethics Committee, Medilink Hospital, Opp. Someshwara Jain Temple, 132 ft Ring Road, Satellite, Ahmedabad, Gujarat 380015, India
6. Human Welfare, Ethical Committee for Human Sciences and Research, Opp. Kama House, Ajmer Road, Jaipur
7. K.R.M Hospital Ethics Committee, 3/92-93 Vijayant khand, Near Chinhut Crossing, Gomtinagar, Lucknow, Uttar Pradesh 226010, India
8. Diacon Hopsital Ethics Committee, 19th Main, 1st Block, Rajajinagar, Bengaluru, Karnataka 560010, India
9. Life Care Hospital Institutional Review Board, #2748/ 2152, M.L.N. Enclave, 16th E Cross, 8th Main, D Block, Next to Corporation Bank, Sahakar Nagar, Bengaluru, Karnataka 560092, India
10. Ramaiah Medical College Ethics Committee, MSRIT Post, MSR Nagar, Bengaluru, Karnataka 560054, India
11. Institutional Ethics Committee, Deenanath Mangeshkar Hospital, Near Mhatre Bridge, Erandawne, Pune, Maharashtra 411004, India
12. Sushruta Hospital Ethics Committee, P. B. Road, Vidyanagar, Hubli, Dharwad, Karnataka, 580021, India
13. Supe Hospital Ethics Committee, Gharapure Ghat Road, Ashok Stambh, Panchavati, Nashik, Maharashtra 422002, India
14. Institutional Ethics Committee of Kovai Diabetes Speciality Centre and Hospital, 15, Vivekanand Road, Ram Nagar-641009, Coimbatore, Tamil Nadu, India
15. Osmania Mediacl College Ethics Committee, Afzal Gunj, Hyderabad, Telangana 500012, India
16. Institutional Ethics Committee, M/S. King George Hopsital, Jagadamba Junction, Visakhapatnam, Andhra Pradesh 530002, India
17. Ethics Committee, S. P. Medical College & Associated Group of Hospitals, S P Medical College Rd, PBM Hospital, Bikaner, Rajasthan 334001, India
18. Institutional Ethics Committee, Gandhi Medical College Hospital,
    Musheerabad, Secunderabad, Telangana 500033, India
19. Institute Ethics Committee, Christian Medical College, CMC Chownk, Brown Road, Ludhiana, Punjab 141008, India
20. Ethics Committee of Bangalore Medical College and Research Institute, K.R. Road, Fort, Bengaluru, Karnataka 560002, India

**Supplementary C: Methods**

**C1. Randomization criteria**

The randomization scheme was produced using a validated system and an interactive voice response system was used to assign treatment at each site. While the treatment allotment was open label, postbaseline HbA_1c_ assessments were blinded to the investigators throughout the treatment period.

The following were the criteria used for additional post run-in inclusion criteria (Randomization criteria):

- FPG of ≤ 140 mg/dL
- 2-h PPG of ≥ 180 mg/dL after STM
- HbA_1c_ ≥ 7% and ≤ 9% (53-75 mmol/mol).
- With appropriate compliance (≥80% compliance with dose and timing of insulin glargine administration, 10/14 time points of SMBG)

**C2. Dosing and dose titration**

After randomization, additional up-titration of study doses were allowed up to a maximum of 8 weeks (maintenance period) for patients persistently lacking PPG control. Permitted maximum dose of Tregopil was 45 mg prior to each meal of the day. Pre-defined rescue criteria based on FPG and/or HbA_1c_ levels indicated when a patient needed to be shifted back to standard of care and not proceed to completion of the study’s 24-weeks treatment period. Up-titration of the already optimized insulin glargine and metformin (+/- OADs) doses were not allowed during the treatment period, but down-titration and reverting to same dose were allowed as a preventive measure to avoid possible hypoglycemia. For hypoglycemia issues, down- and re-titration to the last-highest study dose was permitted for prandial insulins, and titration was supervised by a centralized titration review team.

**C3: Post-hoc analyses of HbA_1c_**

Estimated HbA_1c_ was calculated for all patients at 0, 12 and 24 weeks using the mean daily blood glucose determined from the 9-point SMBG curves using the formula: mean 24 hr blood glucose x 0.0348 + 1.6 (1). The mean daily SMBG based blood glucose was calculated from an overall average of SMBG7 and 9 combined which at baseline is calculated between Visit 3 to 9, at Week 12: Visit 10 to 16 and Week 24: Visit 17 to 19.

**C4. Additional study endpoints**

The following were some of the additional endpoints considered in the study:

- Physical examination
- 12-lead electrocardiogram
- Assessment of adverse events (AEs) and serious AEs (SAEs)
- Symptomatic hypoglycemia SMBG
- 3-point SMBG,
- Standard clinical laboratory evaluations including hematology, clinical chemistry (including fasting lipid profile), and urinalysis.

**Supplementary Table S1:** Demographic and clinical characteristics of study participants at

Baseline

|  | Tregopil | | IAsp  (*N* = 30) | Total population  (*N* = 91) |
| --- | --- | --- | --- | --- |
|  | **30 mg (*N*= 30)** | **45 mg (*N*= 31)** |  |  |
| Gender *n* (%) | | | | |
| Male | 16 (53.3) | 16 (51.6) | 16 (53.3) | 48 (52.7) |
| Female | 14 (46.7) | 15 (48.4) | 14 (46.7) | 43 (47.3) |
| Race *n* (%) | | | | |
| Asian | 30 (100.0) | 31 (100.0) | 30 (100.0) | 91 (100.0) |
| Demographic variables, Mean (SD) | | | | |
| Age (Years) | 50.9 (9.44) | 52.1 (9.49) | 54.5 (8.18) | 52.5 (9.08) |
| Weight (kg) | 68.88 (9.04) | 68.54 (11.55) | 67.76 (9.62) | 68.39 (10.04) |
| Height (cm) | 158.76 (9.26) | 158.84 (7.98) | 158.20 (9.32) | 158.60 (8.77) |
| BMI (kg/m^2^) | 27.39 (3.41) | 27.13 (3.86) | 27.13 (3.66) | 27.22 (3.61) |
| Baseline characteristics, Mean (SD) | | | | |
| Duration of type 2 diabetes (Years) | 7.83 (7.71) | 8.78 (7.86) | 8.12 (6.78) | 8.25 (7.40) |
| HbA_1c_ (%) | 8.10 (0.67) | 8.23 (0.57) | 8.07 (0.69) | 8.13 (0.64) |
| FPG (mg/dL) | 107.2 (30.99) | 109.7 (27.73) | 120.5 (32.20) | 112.4 (30.54) |
| Basal insulin at screening  *n* (%) | 22 (73.3) | 21 (67.7) | 23 (76.7) | 66 (72.5) |

BMI - Body mass index

FPG - Fasting plasma glucose
HbA_1c_ - Hemoglobin A_1c_

IAsp- Insulin Aspart

SD - Standard deviation

| Treatment | Tregopil 30 mg  vs  IAsp | | Tregopil 45 mg  vs  IAsp | | Tregopil 30 mg+45 mg  vs  IAsp | |
| --- | --- | --- | --- | --- | --- | --- |
| PPGE (mg/dL) | **1-h** | **2-h** | **1-h** | **2-h** | **1-h** | **2-h** |
| Mean  ETD  [95% CI]  *P* value | -17.85  [-42.81,7.11]  0.160 | -8.96  [-36.21,18.29]  0.516 | -15.78  [-40.11,8.54]  0.202 | 1.38  [-25.18, 27.94]  0.918 | -16.76  [-38.19 ,4.66]  0.124 | -3.52  [-26.91,19.87]  0.767 |
| CFB  ETD  [95% CI]  *P* value | -45.33  [-71.91, -8.75]  0.001 | -30.22  [-66.65,6.20]  0.103 | -23.62  [-49.53, 2.29]  0.074 | -7.13  [-42.63, 28.38]  0.692 | -33.91  [-56.72, -11.09]  0.004 | -18.07  [-49.33,13.20]  0.255 |
| PPG (mg/dL) | | | | | | |
| Mean  ETD  [95% CI]  *P* value | -1.59  [-32.75, 29.56]  0.920 | 7.30  [-27.07,41.66]  0.675 | -12.14  [-42.51,18.23]  0.431 | 5.02  [-28.48, 38.52]  0.767 | -7.14  [-33.89,19.60]  0.598 | 6.10  [-23.40,35.60]  0.683 |
| CFB  ETD  [95% CI]  *P* value | -11.81  [-43.75, 20.12]  0.466 | 3.30  [-37.18,43.77]  0.872 | -7.31  [-38.44, 23.82]  0.643 | 9.19  [-30.26 ,48.64]  0.646 | -9.44  [-36.86,17.97]  0.497 | 6.40  [-28.35,41.14]  0.716 |

**Supplementary Table S2:** Analyses of 1-h and 2-h Postprandial Plasma Glucose Excursion (PPGE) and Postprandial Plasma Glucose (PPG) by Standard Meal Tolerance (STM) at week 24

CFB - Change From Baseline;

ETD - Estimated treatment difference;

IAsp - Insulin Aspart

PPGE - Postprandial Glucose Excursion;

PPG - Postprandial Glucose;

STM - Standardized Test Meal;

SD - Standard Deviation;

The treatment differences mentioned here are the LSMD (Least square mean difference) values of mean and CFB of PPGE and PPG from STM at week 24.

**Supplementary Table S3:** Summary of 1-h and 2-h PPGE and PPG from 9-point SMBG at week 24

| Time-points | Tregopil 30 mg  (*N* = 30)* | Tregopil 45 mg  (*N* = 31)# | IAsp  (*N* = 30)¥ |
| --- | --- | --- | --- |
| PPGE (mg/dL) | **Mean (SD)** | **Mean (SD)** | **Mean (SD)** |
| 1-h postbreakfast | 47.79 (44.84) | 52.59 (40.40) | 62.07 (43.17) |
| 2-h postbreakfast | 47.19 (46.22) | 59.78 (48.49) | 60.29 (45.75) |
| 1-h postlunch | 24.12 (46.07) | 31.41 (56.10) | 41.31 (48.97) |
| 2-h postlunch | 44.12 (41.51) | 46.89 (59.78) | 31.78 (48.73) |
| 1-h postdinner | 18.11 (56.34) | 24.77 (40.11) | 22.10 (55.24) |
| 2-h postdinner | 40.69 (62.23) | 44.56 (55.40) | 20.12 (69.58) |
| PPG (mg/dL) | | | |
| 1-h postbreakfast | 178.75 (47.45) | 182.68 (46.52) | 180.79 (57.65) |
| 2-h postbreakfast | 178.91 (51.38) | 186.35 (51.41) | 179.98 (54.98) |
| 1-h postlunch | 177.40 (40.38) | 180.37 (50.39) | 174.69 (45.51) |
| 2-h postlunch | 199.44 (51.68) | 195.85 (50.77) | 170.36 (41.79) |
| 1-h postdinner | 184.26 (49.83) | 184.77 (47.12) | 164.71 (49.74) |
| 2-h postdinner | 202.33 (50.98) | 199.38 (45.60) | 162.72 (56.22) |

IAsp- Insulin Aspart

PPGE - Postprandial glucose excursion

SMBG - Self monitored blood glucose

SD - Standard deviation

**n* = 26 for 1-h postbreakfast, 2-h postlunch and 2-h postdinner; *n* = 27 for 2-h postbreakfast, 1-h postdinner; *n* = 25 for 1-h postlunch for PPG and PPGE in Tregopil 30 mg group

**#***n* = 27 for 1-h and 2-h postbreakfast; *n* = 28 for 1-h and 2-h postlunch, and 1-h postdinner; *n* = 25 for 2-h postdinner for PPG and PPGE in Tregopil 45mg group

**¥***n* = 27 for 1-h postbreakfast; *n* = 29 for 2-h postbreakfast, 2-h postlunch and 1-h and 2-h postdinner; *n* = 26 for 1-h postlunch for PPG and PPGE in IAsp group

**Supplementary Table S4:** Analyses of 1-h and 2-h PPGE and PPG by SMBG at week 24

| Time point |  | Tregopil 30 mg vs IAsp | | Tregopil 45 mg vs IAsp | | Tregopil 30 mg +45 mg vs IAsp | |
| --- | --- | --- | --- | --- | --- | --- | --- |
| PPGE (mg/dL) |  | **1-h** | **2-h** | **1-h** | **2-h** | **1-h** | **2-h** |
| Breakfast | **Mean**  ETD [95%CI]  *P* value | -14.29  [-37.45, 8.88]  0.225 | -13.11  [-37.98, 11.77]  0.299 | -9.48  [-32.43, 13.46]  0.415 | -0.52  [-25.39, 24.36]  0.967 | -11.84  [-31.77, 8.10]  0.242 | -6.81  [-28.23, 14.60]  0.530 |
|  | **CFB**  ETD [95%CI]  *P* value | -16.77  [-46.19, 12.65]  0.262 | 8.53  [-19.09, 36.14]  0.542 | -5.19  [-34.33, 23.96]  0.725 | 7.42  [-20.20, 35.03]  0.596 | -10.87  [-36.19, 14.45]  0.397 | 7.97  [-15.80, 31.74]  0.508 |
| Lunch | **Mean**  ETD [95%CI]  *P* value | -17.19  [-45.44, 11.06]  0.231 | 12.34  [-14.90, 39.58]  0.372 | -9.90  [-37.37, 17.57]  0.477 | 15.12  [-11.61, 41.84)  0.265 | -13.34  [-37.48, 10.81]  0.277 | 13.78  [-9.44, 37.00]  0.243 |
|  | **CFB**  ETD [95%CI]  *P* value | -24.55  [-53.36, 4.26]  0.094 | 0.38  [-28.99, 29.74]  0.980 | 3.02  [-24.99, 31.03]  0.831 | 8.81  [-20.00, 37.62]  0.546 | -9.99  [-34.61, 14.64]  0.424 | 4.75  [-20.28, 29.78]  0.708 |
| Dinner | **Mean**  ETD [95%CI]  *P* value | -3.99  [-30.46, 22.47]  0.766 | -20.57  [12.17, 53.32]  0.216 | 2.66  [-23.56, 28.89]  0.841 | 24.44  [-8.65, 57.53]  0.146 | -0.60  [-23.32, 22.11]  0.958 | 22.47  [-5.73, 50.67]  0.117 |
|  | **CFB**  ETD [95%CI]  *P* value | -14.26  [-48.73, 20.20]  0.415 | 12.48  [-26.37, 51.33]  0.526 | -16.50  [-50.65, 17.64]  0.341 | 3.67  [-35.58, 42.93]  0.853 | -15.40  [-44.98, 14.17]  0.305 | 8.16  [-25.29, 41.62]  0.630 |
| PPG (mg/dL) |  |  | | | | | |
| Breakfast | Mean  ETD [95%CI]  *P* value | -2.05  [-28.77, 24.68]  0.880 | -1.08  [-28.60, 26.45]  0.939 | 1.89  [-24.58, 28.36]  0.888 | 6.37  [-21.16, 33.90]  0.648 | -0.04  [-23.04, 22.96]  0.997 | 2.65  [-21.05, 26.34]  0.825 |
|  | CFB  ETD [95%CI]  *P* value | 2.99  [-30.76, 36.74]  0.861 | 22.56  [-8.15, 53.27]  0.149 | 14.64  [-18.79, 48.07]  0.388 | 23.58  [-7.13, 54.29]  0.131 | 8.92  [-20.12, 37.96]  0.544 | 23.07  [-3.36, 49.50]  0.087 |
| Lunch | Mean  ETD [95%CI]  *P* value | 2.71  [-22.62, 28.04]  0.833 | 29.08  [2.80, 55.36]  0.030 | 5.68  [-18.94, 30.31]  0.649 | 25.50  [-0.29, 51.28]  0.053 | 4.28  [-17.37, 25.93]  0.696 | 27.22  [4.82, 49.62]  0.018 |
|  | CFB  ETD [95%CI]  *P* value | 0.52  [-30.00, 31.04]  0.973 | 19.52  [-9.92, 48.96]  0.192 | 23.09  [-6.59, 52.77]  0.126 | 27.42  [-1.46, 56.30]  0.063 | 12.44  [-13.65, 38.53]  0.347 | 23.62  [-1.48, 48.71]  0.065 |
| Dinner | Mean  ETD [95%CI]  *P* value | 19.55  [-6.13, 45.24]  0.135 | 39.60  [12.84, 66.36]  0.004 | 20.06  [-5.38, 45.51]  0.121 | 36.66  [9.61, 63.70]  0.008 | 19.81  [-2.23, 41.85]  0.078 | 38.16  [15.11, 61.20]  0.001 |
|  | CFB  ETD [95%CI]  *P* value | 12.64  [-20.13, 45.40]  0.447 | 39.84  [6.44, 73.24]  0.020 | 15.18  [-17.28, 47.64]  0.357 | 34.20  [0.45, 67.95]  0.047 | 13.93  [-14.18, 42.05]  0.329 | 37.07  [8.31, 65.83]  0.012 |

CFB - Change From Baseline; CI - Confidence interval; ETD - Estimated treatment difference; IAsp - Insulin Aspart; PPGE - Postprandial Glucose Excursion; PPG - Postprandial Glucose; SMBG - Self monitored blood glucose

**Supplementary Table S5**: Mean measured HbA_1c_ and estimated A_1c_ (eA1c) at baseline and at week 24

| **Visit** | | **Tregopil 30 mg**  **(*n* = 30)*** | **Tregopil 45 mg**  **(*n* = 31)**^#^ | **IAsp**  **(*n* = 30)**$ |
| --- | --- | --- | --- | --- |
|  |  | **Mean (SD)** | **Mean (SD)** | **Mean (SD)** |
| **Baseline** | **Measured A_1c_** | 8.10 (0.67) | 8.23 (0.58) | 8.07 (0.70) |
|  | **Mean Glucose (9-point SMBG)** | 183.47 (28.81) | 186.60 (28.59) | 185.50 (30.42) |
|  | **Estimated A_1c_** | 7.98 (1.00) | 8.09 (1.00) | 8.06 (1.06) |
| **Week 24** | **Measured A_1c_** | 8.21 (0.99) | 8.39 (1.16) | 7.29 (0.79) |
|  | **Mean glucose (9-point SMBG)** | 176.28 (36.09) | 177.34 (32.19) | 158.85 (27.04) |
|  | **Estimated A_1c_** | 7.73 (1.26) | 7.77 (1.12) | 7.13 (0.94) |

N = Total number of randomized patients (total number of patients in the Intent-to-Treat analysis set in each treatment arm)

For Mean glucose 9-point curve ***number of patients (n = 28) in Tregopil 30 mg group at weeks 24; *^#^*number of patients (n = 29) in Tregopil 45 mg group at weeks 24; ^$^number of patients (n = 29) in IAsp group at weeks 24

For Measured A1c ***number of patients (n = 29) in Tregopil 30 mg group at weeks 12 and weeks 24; *^#^*number of patients (n = 30) in Tregopil 45 mg group at weeks 12 and weeks 24; ^$^number of patients (n = 29) in IAsp group at weeks 12 and weeks 24.

HbA_1c_: Glycated hemoglobin; Estimated A_1c_ = Mean Glucose(mg/dL) *0.0348+ 1.6

Baseline was defined as the last observed value of a parameter before first intake of trial medication. Change from baseline (CFB) was calculated as the difference between value of interest and corresponding baseline value.

**Supplementary Table S6:** Incidences of hypoglycemic events and TEAEs during the 24-week study period

|  | Tregopil 30 mg  (*N* = 30) | | Tregopil 45 mg  (*N* = 31) | | IAsp  (*N* = 30) | |
| --- | --- | --- | --- | --- | --- | --- |
|  | ***n* (%)** | **No. of events**  **(Rates)** | ***n* (%)** | **No. of events (Rates** | ***n* (%)** | **No. of events**  **(Rates)** |
| Incidence of hypoglycemic events | | | | | | |
| Postbreakfast | 18 (60.0) | 54 | 19 (61.3) | 49 | 20 (66.7) | 70 |
| Postlunch | 15 (50.0) | 23 | 13 (41.9) | 22 | 20 (66.7) | 44 |
| Postdinner | 12 (40.0) | 19 | 13 (41.9) | 17 | 18 (60.0) | 46 |
| No. of patients with clinically significant hypoglycemia (level 2) | 16 (53.3) | 38 (282.9) | 13 (41.9) | 27 (193.3) | 17 (56.7) | 47 (346.3) |
| No. of patients with at least one TEAE | 6 (20.0) | 7 | 5 (16.1) | 15 | 4 (13.3) | 7 |

IAsp - Insulin Aspart

TEAEs - Treatment Emergent Adverse Events

**Supplementary Figure S1:** Patient disposition

Enrollment

Randomized in 1:1:1 ratio (*N* = 91)

Analyzed in week 12 (*n* = 29); week 24 (*n* = 27)

Analyzed in week 12 (*n* = 30); week 24 (n = 29)

Analyzed in week 12 (*n* = 29); week 24 (*n* = 29)

Assessed for eligibility (*N* = 269)

Excluded (*n* = 178)

- Not meeting inclusion criteria (*n*= 126)
- Declined patients (due to run-in failures) (*n* = 52)

Allocation

Follow - up

Analysis

Tregopil 30 mg group (*n* = 30)

Tregopil 45 mg group (*n* = 31)

Insulin Aspart (4 U/meal) group (*n* = 30)

Lost to follow-up (*n* = 3)

- adverse event, *n* =1
- withdrawal of consent, *n* = 1
- rescue criteria, *n* = 1

Lost to follow-up (*n* = 2)

- withdrawal of consent, *n* = 1
- rescue criteria, *n* = 1

Lost to follow-up (*n* = 1)

- withdrawal of consent, *n* = 1

**Supplementary Figure S2:** Study design

Screening period (3 weeks)

Run-in period (8 weeks)

Treatment period (24 weeks). Assessed the efficacy, safety and treatment satisfaction at end of treatment

Safety follow-up period (2 weeks)

Screened criteria:

- HbA_1c_ 7.5% -10%
- On stable dose of metformin ± oral antidiabetics drugs ± basal insulin

Run in period:

- Initial 4 weeks of insulin glargine dose titration period
- Subsequent 4 weeks of dose stabilization period
- Metformin was given once a day or twice a day or as applicable as a stable dose from screening

Patients who met the following eligible criteria at week 8 were randomized to 1:1:1 ratio to receive Tregopil 30 mg, 45 mg, and insulin Aspart:

- HbA_1c_ ≥ 7% and ≤ 9%
- Fasting plasma glucose ≤ 140 mg/dL, 2-h PPG ≥ 180 mg/dL after standardized test meal.

However, insulin glargine and metformin were not uptitrated in the treatment period

**Insulin Aspart grou**p

- The dose was started at 4 units/meal TID (before 5 min of each meal) through a subcutaneous route
- In a first 4 weeks of treatment, the dose of Tregopil was titrated based on the titration algorithms and meal-specific average of the 9-point SMBG levels
- The dose was stabilized and optimized in rest of the treatment period up to 24 weeks
- In patients with persistent lack of PPG control during the first 4 weeks, additional titration was permitted up to a maximum of 8 weeks in all the three groups.

**45 mg of Tregopil group**

- The dose was started at 45 mg TID (2 tablets of 15 mg each administered 10 ± 2 min before the 3 major meals)
- In a first 4 weeks of treatment, the dose of Tregopil was titrated based on the titration algorithms and meal-specific average of the 9-point SMBG levels
- The dose was stabilized and optimized in rest of the treatment period up to 24 weeks

**30 mg of Tregopil group**

- The dose was started at 30 mg TID (2 tablets of 15 mg each administered 10 ± 2 min before the 3 major meals)
- In a first 4 weeks of treatment, the dose of Tregopil was titrated based on the titration algorithms and meal-specific average of the 9-point SMBG levels
- The dose was stabilized and optimized in rest of the treatment period up to 24 weeks

End of the study recorded the adverse events and hypoglycemic events through phone call. If occurrence of any adverse events and hypoglycemic events indicated clinic visit, this was converted into a clinic visit, as per investigator discretion.

For hypoglycemia concerns, down- and re-up titration to the last-highest study dose was allowed for the prandial insulins and titration was monitored by a centralized titration review team.

TID-Thrice in a day; PPG- Postprandial Glucose; SMBG - Self monitored blood glucose

**Supplementary Figure S3:** Percentage of patients achieving 2-h PPG level (<140 mg/dL) after STM and/or measured by 9-point SMBG at week 24

% of patients

|  | Tregopil 30 mg | Tregopil 45mg | IAsp |
| --- | --- | --- | --- |
| STM (<140 mg/dL) | 21.4 | 26.7 | 17.2 |
| SMBG (<140 mg/dL) | 25 | 23.3 | 17.2 |

PPG - Postprandial glucose; STM – Standardized test meal; SMBG - Self monitored blood glucose; IAsp- Insulin Aspart

**Supplementary Figure S4:** CFB in PPG levels (1-h and 2-h) from 9-point SMBG at week 24 following each meal of the day

Treatment differences of CFB

| Postmeal time points (h) | Tregopil 30 mg vs IAsp | Tregopil 45 mg vs IAsp |
| --- | --- | --- |
| Breakfast 1 h | 2.99 | 14.64 |
| Breakfast 2 h | 22.56 | 23.58 |
| Lunch 1 h | 0.52 | 23.09 |
| Lunch 2 h | 19.52 | 27.42 |
| Dinner 1 h | 12.64 | 15.18 |
| Dinner 2 h | 39.84 | 34.2 |

PPG - Postprandial glucose; CFB - Change from baseline; SMBG - Self monitored blood glucose; IAsp- Insulin Aspart
